# EXPLORING MALE INVOLVEMENT IN CONTRACEPTIVE DECISION-MAKING AND SUPPORT FOR LONG-ACTING REVERSIBLE CONTRACEPTIVES AMONG COUPLES IN KAMPALA, UGANDA

**DOI:** 10.64898/2026.03.11.26348000

**Authors:** Brian Nyasulu, Nicholas Ngomi, Arineitwe Ronald Kibonire, Anumolu Goparaju

**Author notes:** **Corresponding author:** Brian Nyasulu.

## Abstract

Male involvement in family planning remains a critical yet underexplored factor influencing contraceptive uptake and decision-making support for long-acting reversible contraceptives (LARCs) in Kampala, Uganda. This mixed-methods study assessed male participation in decision-making and support for long-acting reversible contraceptives among couples in Kampala. The study involved 362 male participants who completed structured questionnaires, six focus group discussions (FGDs) conducted with both men and women, and five key informant interviews (KIIs) with healthcare providers. Ethical approval was obtained from the Kampala International University Research Ethics Committee and the Uganda National Council for Science and Technology.

Quantitative findings revealed that 96.4% of men reported that their partners discussed contraception with healthcare providers (χ² = 31.366, p < 0.001), yet only 9.7% of men accompanied their partners to clinics. Male support for family planning was primarily financial (60.2%), while joint decision-making on LARCs was reported by only 38.7% (χ² = 2.776, p = 0.596). Key determinants of male involvement included marital status, education level, and number of children. However, cultural norms (χ² = 42.813, p < 0.000) and religious beliefs (χ² = 29.402, p < 0.021) were identified as significant barriers to male participation.

Qualitative findings from FGDs and KIIs echoed the limited involvement of men in family planning services, attributing this to entrenched gender norms, misconceptions about contraceptives, and the perception that reproductive health services are primarily for women. Although 44.2% of participants indicated that men’s concerns were addressed during consultations, 77.3% reported limited availability of couple-focused counseling (χ² = 6.294, p = 0.178).

The study concludes that male involvement in family planning decision-making and support for LARCs remains low, largely due to socio-cultural barriers and limited male-friendly services. The study recommends strengthening male engagement strategies by training health workers on inclusive counseling, involving community and religious leaders in awareness campaigns, and promoting male-friendly and couple-centered reproductive health services through community-based platforms.

## Introduction

Family planning is a critical component of reproductive health, contributing significantly to improved maternal and child health outcomes, reduced unintended pregnancies, and enhanced gender equality (UNFPA, 2021). Despite the well-documented benefits of family planning, male involvement in these programs remains limited, particularly concerning the use of long-acting reversible contraceptive (LARC) methods. Traditionally, family planning initiatives have predominantly targeted women, often overlooking the influential role men play in reproductive decision-making within many societies(Takyi et al., 2023). This gender imbalance hampers the effectiveness of family planning programs and underscores the need to actively engage men as partners in reproductive health.

Globally, healthcare providers are recognized as pivotal actors in promoting male participation in family planning by creating inclusive, male-friendly environments and providing education to dispel myths and misconceptions about contraceptives(Mwaisaka et al., 2020). Research from a variety of settings shows how crucial provider attitudes, communication abilities, and collaborative decision-making are in encouraging male participation, which improves reproductive health outcomes (UNFPA, 2021). Healthcare professionals in Tanzania, for example, expressed pleasure with the rise in male involvement but also pointed out that service delivery is constrained by resource and infrastructure shortages(Msovela et al., 2022).

Data from Uganda’s National Family Planning Costed Implementation Plan (2020/21–2024/25) shows that male participation in family planning is alarmingly low, with a prevalence rate of just 28.4% (UNFPA, 2021). Similarly, the 2024 Uganda Bureau of Statistics report indicates that only 38% of married women aged 15 to 49 use any contraceptive method (UBOS, 2024).These statistics underscore a critical gap in family planning service delivery and uptake, particularly among men, which undermines national efforts to reduce unintended pregnancies and improve maternal and child health.

Additional obstacles that healthcare workers in Sub-Saharan Africa must overcome include cultural norms that discourage male involvement, a lack of resources, and poor training (World Health Organization, 2018). To promote male involvement through outreach initiatives and education, providers work to integrate culturally appropriate approaches and partner with community leaders(Kriel et al., 2023). Male-centered care and community-based programs are the main focus of current efforts in Uganda to raise male awareness and accessibility to family planning services, especially in underserved and rural areas(Wambete et al., 2022). However, obstacles such enduring gender stereotypes, false information, and restrictions on the health system still prevent males from participating, particularly in urban and peri-urban areas like Kampala(Alupo et al., 2020).

Despite these revelations, there is still a great deal of unanswered questions about the precise role that healthcare professionals play in encouraging male involvement in LARC usage, especially in Ugandan urban slum areas. Male engagement is typically aggregated in research without a detailed analysis of views or support for LARC procedures(Jonas et al., 2020). Furthermore, in these settings, provider biases, insufficient infrastructure, and cultural challenges have not yet been thoroughly investigated(Alupo et al., 2020). To improve male participation in family planning and reproductive health outcomes, it is essential to address these gaps and develop targeted interventions that consider the unique socioeconomic and cultural dynamics of urban populations.

## Methodology

This study employed a mixed-methods cross-sectional design to examine male involvement in long-acting reversible contraceptive (LARC) decision-making among couples in Kampala, Uganda. **Participant recruitment for the study was conducted between 24/03/2025 and 30/09/2025** in Makindye Division and Kisugu Health Centre III. Quantitative data were collected through structured questionnaires administered to 362 men aged 18 years and above.

Qualitative data were obtained through semi-structured interviews with women and their male partners, six focus group discussions (FGDs), and Five key informant interviews (KIIs) with health workers involved in family planning service delivery.

Quantitative data were analyzed using the Statistical Package for the Social Sciences (SPSS), while qualitative data were analyzed thematically using NVivo software. The structured questionnaire and focus group discussion guide were specifically developed for this study and are provided in English as Supplementary File 1.

Ethical approval was obtained from the **Kampala International University Research Ethics Committee** (KIU-REC) (Approval No. KIU-2024-563) and the **Uganda National Council for Science and Technology** (UNCST) (Approval No. HS5778ES). All study procedures adhered to ethical standards, including obtaining written informed consent, ensuring confidentiality, and maintaining cultural sensitivity throughout the research process.

## Results

The study examined key socio-demographic and belief-related factors influencing male involvement in the use and decision-making around long-acting reversible contraceptives (LARCs) among 362 respondents in Kampala, Uganda.

Marital status was significantly associated with male involvement (χ² = 35.627, *p* = 0.001), with the majority of respondents being married (86.2%), followed by single (6.1%) and cohabiting individuals (4.4%).

Education level also showed a significant association (χ² = 44.289, *p* = 0.002), with most participants having secondary (43.4%) or diploma/certificate education (30.1%).

The number of children was another significant factor (χ² = 38.428, *p* = 0.004), where most respondents had between three to four children (48.1%) or one to two children (38.7%).

Regarding barriers to male involvement in LARCs, fear of side effects (35.7%) and partner preference (30%) were the most cited reasons, followed by religious beliefs (21.1%) and cultural beliefs (8.6%). However, this variable was not statistically significant (χ² = 21.053, *p* = 0.636).

Cultural norms had a strong influence on involvement, with 73% agreeing and 8.8% strongly agreeing, and this was statistically significant (χ² = 42.813, *p* < 0.001).

Similarly, religious beliefs were significantly associated (χ² = 29.402, *p* = 0.021), with 40.3% agreeing and 6.1% strongly agreeing that religion influenced their decisions about LARCs.

Qualitative findings from focus group discussions reinforced these quantitative results. Married men in Kampala shared diverse perspectives on their roles in family planning. One participant in his 30s expressed a sense of responsibility in reproductive decision-making, stating

“*In my marriage, I feel responsible for every decision that concerns our family, especially the number of children we plan to have. My wife and I often talk about family planning, and we both agreed to go for a long-term method after our third child*.” (FGD, Woman 1)

Another participant, a secondary school teacher aged 36–40 years with a diploma in education, highlighted the influence of awareness and education

“*I learned about LARC methods during a health outreach session at our local center. Before that, I used to think family planning was just a woman’s issue. But now I know it’s something we should decide on together. Education helped me understand how important this is, not just for her health but for our future as a family*.” (FGD, Man 2)

Participants consistently emphasized that education, awareness, and financial stability promote joint decision-making, while misinformation, cultural taboos, and fear of stigma remain barriers. Many men expressed the need for accurate information directly from healthcare providers rather than relying on second-hand sources. Strengthening male-focused health education and integrating men into family planning discussions at the facility and community level could therefore enhance shared decision-making and increase support for LARC use among couples.

The quantitative and qualitative findings from this study are strongly complementary and together provide a deeper understanding of male involvement in long-acting reversible contraceptive (LARC) decision-making among couples in Kampala.

Quantitative findings showed that while nearly all respondents (96.4%) had discussed contraception with a healthcare provider a statistically significant result (χ² = 31.366, p < 0.001) direct male involvement in family planning discussions remained limited. Specifically, 77.3% of men reported that healthcare providers had never directly involved them in contraceptive discussions with their partners. This numerical gap between high provider contact and low male inclusion is clearly explained by qualitative evidence. Men described clinic encounters in which family planning conversations were directed primarily toward women, leaving men feeling sidelined or perceived merely as supporters rather than active decision-makers.

This disconnect is illustrated by a male participant who stated

*“When I go to the clinic, they assume it’s my wife’s decision. I also have concerns, but many men fear being judged or seen as less of a man if they talk about contraception.” (FGD, Man 5)*

Similarly, although 44.2% of respondents agreed that healthcare providers adequately address men’s concerns about LARC, and only 23.8% disagreed, this variation was not statistically significant (χ² = 17.663, p = 0.344). Qualitative findings help interpret this lack of statistical significance by revealing wide variability in men’s personal experiences with providers. Some men reported receiving clear information and reassurance, while others expressed persistent fears related to infertility, sexual performance, and long-term side effects concerns that were not always adequately addressed during consultations.

Healthcare providers themselves acknowledged these challenges. A reproductive health officer explained:

*“Many men believe long-term methods cause permanent infertility or affect sexual performance. There is also stigma because family planning is seen as a woman’s domain.” (KII, Reproductive Health Officer 1)*

The quantitative data further showed strong support for enhanced education and outreach, with over 90% of respondents agreeing or strongly agreeing that increased provider-led education could encourage male involvement. Although this finding was not statistically significant (χ² = 12.309, p = 0.421), qualitative data clarify that this consensus reflects a shared recognition of unmet informational needs rather than differences across respondent groups. Participants consistently emphasized the importance of male-focused messaging, joint counseling sessions, and visible male role models to normalize men’s participation in family planning.

As one male participant noted:

*“Healthcare providers need to speak directly to men. When we see other men involved, it feels more normal. It’s not just a woman’s responsibility.” (FGD, Woman 4)*

Finally, healthcare providers highlighted strategies that directly address the gaps identified in the quantitative findings. These include intentionally inviting men into consultations, encouraging couple-based counseling, addressing misconceptions about LARC safety and reversibility, and engaging community leaders to challenge entrenched gender norms.

*“We try to include both partners and explain that family planning is a shared responsibility. Creating a comfortable environment for men is essential.” (KII, Family Planning Coordinator 1)*

## Discussion

The study in Figure 1 found that most men were familiar with injections (53.6%), followed by implants (28.2%) and intrauterine devices (12.4%). Only a few knew about birth control pills (4.4%) and natural methods like withdrawal (1.4%). This shows that awareness of long-acting reversible contraceptives (LARCs) is still limited to a few methods. The high familiarity with injections may be because they are more commonly promoted and easier to access. These findings are similar to those (Ontiri et al., 2021) have observed that women often choose methods that are discreet and easy to use without needing much discussion with their partners. Low awareness of methods like IUDs and implants may affect how couples make decisions and reduce the chances of male involvement. To improve this, both men and women need better education on all types of family planning methods to support shared decision-making.

**Figure 1.**
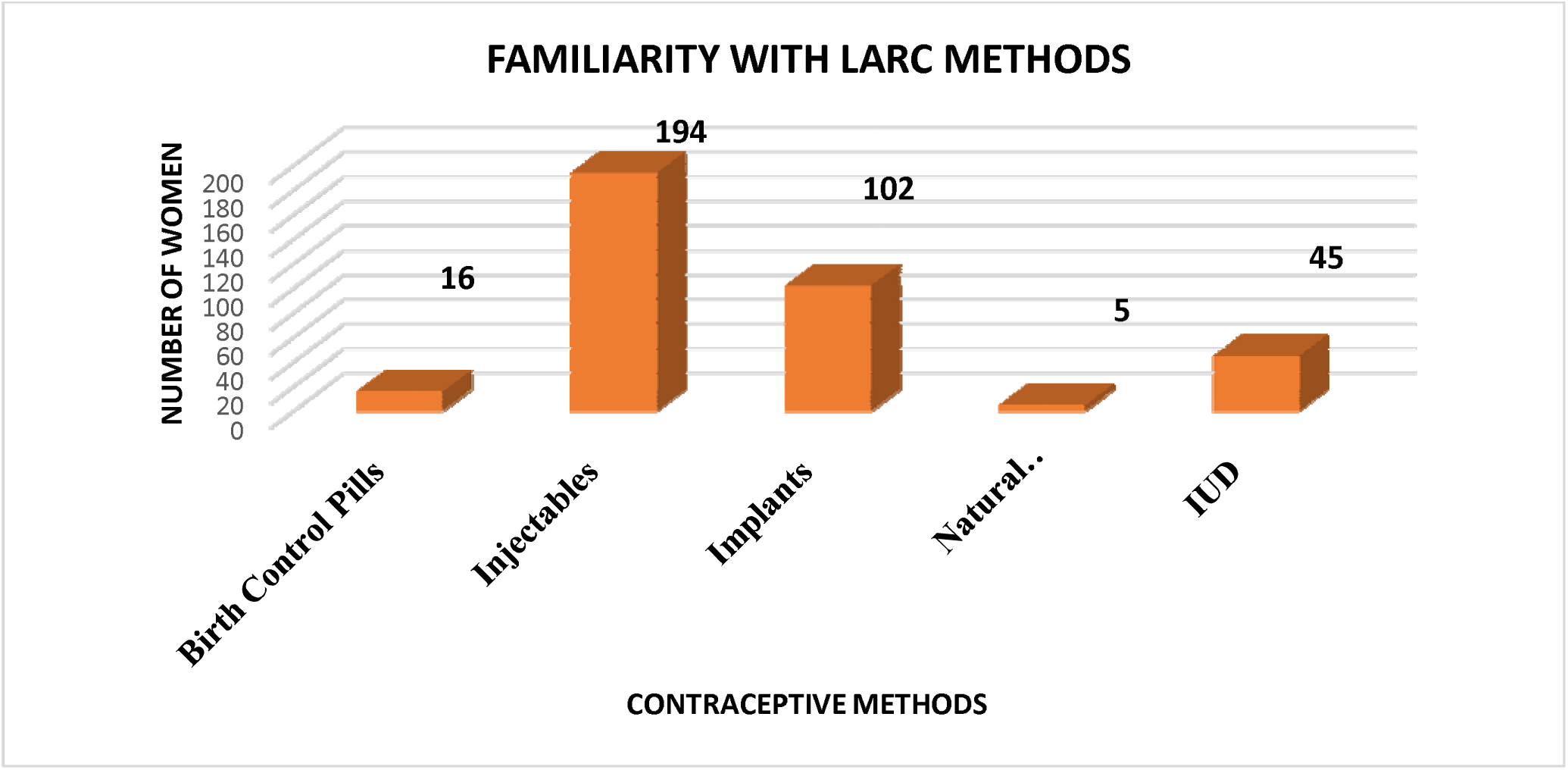
Familiarity with LARC Methods Among Female Respondents. The findings show that among female respondents, injections were the most recognized long-acting reversible contraceptive (LARC) method (53.6%), followed by implants (28.2%) and intrauterine devices (12.4%). Fewer women were familiar with birth control pills (4.4%) or natural methods like withdrawal (1.4%), as shown in Figure (1) below. This indicates varying levels of awareness, which could impact contraceptive choices and male involvement in their use.

The study in Table 1 showed that most respondents believe contraceptive use improves relationships, with 74.3% agreeing that it has a positive effect. This supports findings from (Hernandez et al., 2022), who found that when couples openly discuss contraceptives, it leads to fewer arguments and more satisfaction in relationships. The chi-square test also showed a strong link between positive views on contraception and better relationship quality (χ² = 41.131, p = 0.001). This highlights the importance of involving both partners in contraceptive decisions, rather than leaving the responsibility to women alone.

**Table 1:**
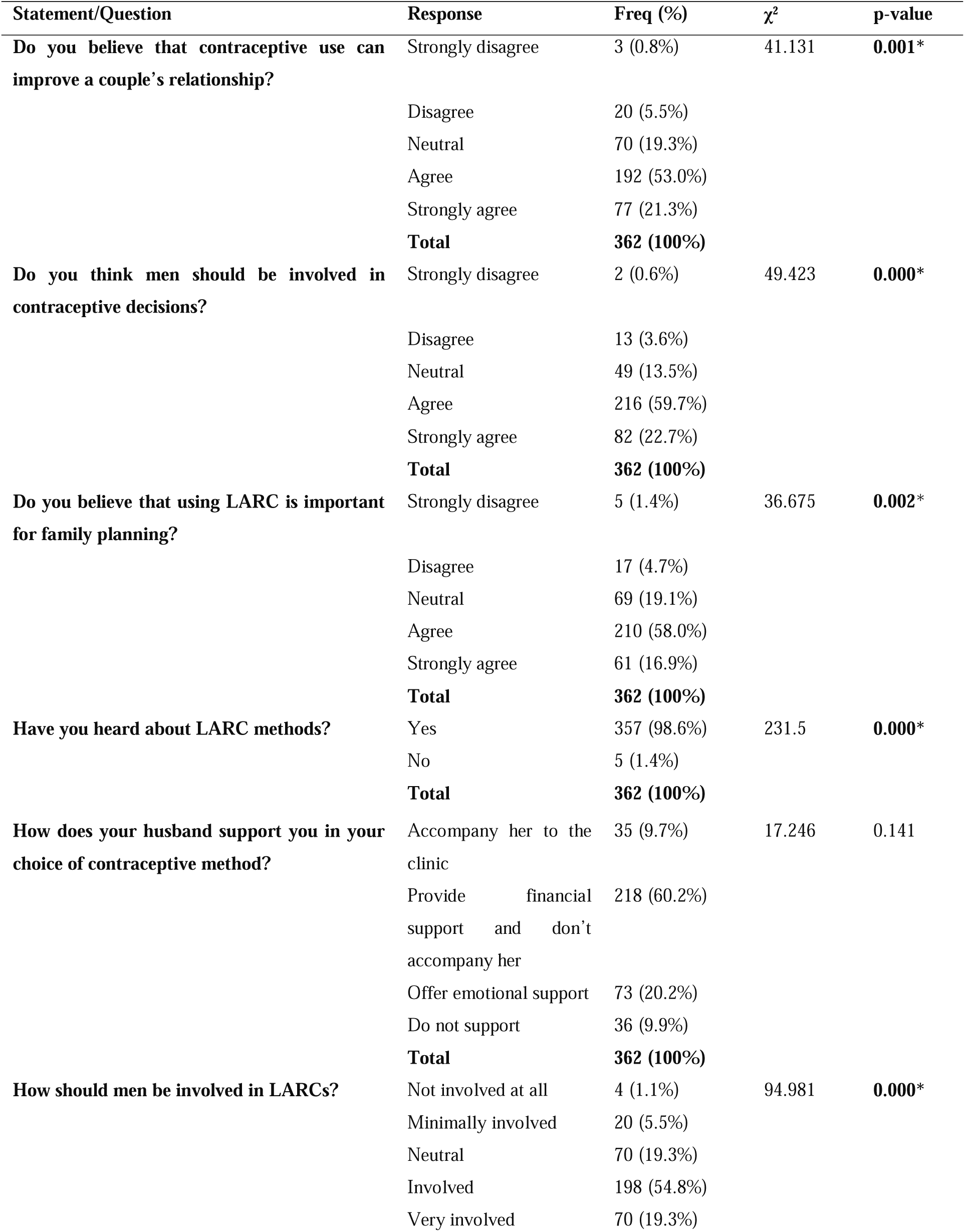
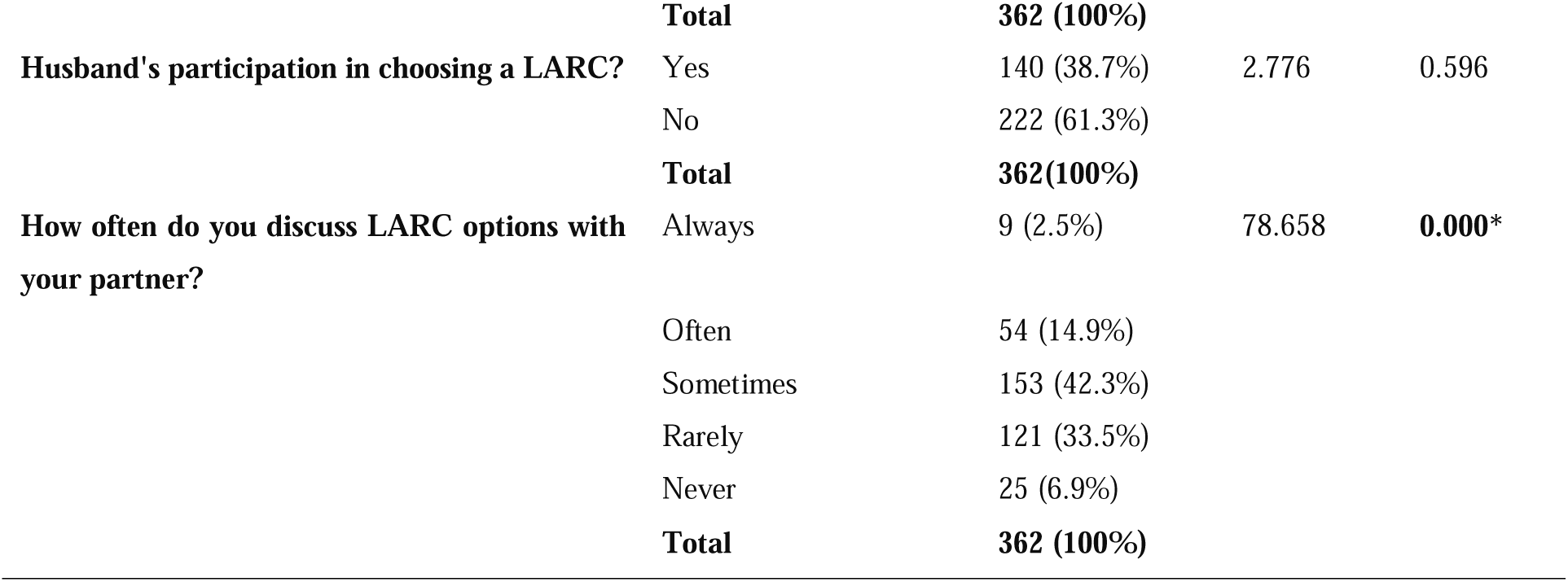
Chi-Square analysis on the extent of male involvement in decision-making about long-acting reversible contraceptives (LARC) among couples in Kampala. The study looked at how men are involved in family planning, especially in the use of long-acting reversible contraceptives (LARCs), among 362 participants in Kampala, Uganda. Most respondents (74.3%) agreed that using contraceptives can improve a couple’s relationship (χ*² = 41.131, p = 0.001*), and 82.4% believed men should be involved in making contraceptive decisions (χ*² = 49.423, p < 0.001*). About 74.9% said LARCs are important for family planning (χ*² = 36.675, p = 0.002*). Awareness of LARC methods was very high, with 98.6% having heard of them (χ*² = 231.500, p < 0.001*). However, only 38.7% of couples had chosen a method together, which was not statistically significant (χ*² = 2.776, p = 0.596*). When it came to support, 60.2% of men said their partners gave financial help but did not go with them to clinics. Only 9.7% said their partners accompanied them, which also was not statistically significant (χ*² = 17.246, p = 0.141*). Even though 74.1% believed men should be involved in family planning (χ*² = 94.981, p < 0.001*), only 17.4% said they often or always talked about LARC options with their partners (χ*² = 78.658, p < 0.001*). This shows that while many support male involvement, actual participation and communication remain low.

| Statement/Question | Response | Freq (%) | $\chi^2$ | p-value |
| --- | --- | --- | --- | --- |
| **Do you believe that contraceptive use can improve a couple's relationship?** | Strongly disagree | 3 (0.8%) | 41.131 | **0.001\*** |
|  | Disagree | 20 (5.5%) |  |  |
|  | Neutral | 70 (19.3%) |  |  |
|  | Agree | 192 (53.0%) |  |  |
|  | Strongly agree | 77 (21.3%) |  |  |
|  | **Total** | **362 (100%)** |  |  |
| **Do you think men should be involved in contraceptive decisions?** | Strongly disagree | 2 (0.6%) | 49.423 | **0.000\*** |
|  | Disagree | 13 (3.6%) |  |  |
|  | Neutral | 49 (13.5%) |  |  |
|  | Agree | 216 (59.7%) |  |  |
|  | Strongly agree | 82 (22.7%) |  |  |
|  | **Total** | **362 (100%)** |  |  |
| **Do you believe that using LARC is important for family planning?** | Strongly disagree | 5 (1.4%) | 36.675 | **0.002\*** |
|  | Disagree | 17 (4.7%) |  |  |
|  | Neutral | 69 (19.1%) |  |  |
|  | Agree | 210 (58.0%) |  |  |
|  | Strongly agree | 61 (16.9%) |  |  |
|  | **Total** | **362 (100%)** |  |  |
| **Have you heard about LARC methods?** | Yes | 357 (98.6%) | 231.5 | **0.000\*** |
|  | No | 5 (1.4%) |  |  |
|  | **Total** | **362 (100%)** |  |  |
| **How does your husband support you in your choice of contraceptive method?** | Accompany her to the clinic | 35 (9.7%) | 17.246 | 0.141 |
|  | Provide financial support and don't accompany her | 218 (60.2%) |  |  |
|  | Offer emotional support | 73 (20.2%) |  |  |
|  | Do not support | 36 (9.9%) |  |  |
|  | **Total** | **362 (100%)** |  |  |
| **How should men be involved in LARCs?** | Not involved at all | 4 (1.1%) | 94.981 | **0.000\*** |
|  | Minimally involved | 20 (5.5%) |  |  |
|  | Neutral | 70 (19.3%) |  |  |
|  | Involved | 198 (54.8%) |  |  |
|  | Very involved | 70 (19.3%) |  |  |
|  | **Total** | **362 (100%)** |  |  |
| **Husband's participation in choosing a LARC?** | Yes | 140 (38.7%) | 2.776 | 0.596 |
|  | No | 222 (61.3%) |  |  |
|  | **Total** | **362(100%)** |  |  |
| **How often do you discuss LARC options with your partner?** | Always | 9 (2.5%) | 78.658 | **0.000\*** |
|  | Often | 54 (14.9%) |  |  |
|  | Sometimes | 153 (42.3%) |  |  |
|  | Rarely | 121 (33.5%) |  |  |
|  | Never | 25 (6.9%) |  |  |
|  | **Total** | **362 (100%)** |  |  |

A major finding of the study was that many people support men being involved in contraceptive decisions. About 82.4% of respondents either agreed or strongly agreed that male participation is important. This matches research by (Wambete et al., 2024), who found that male involvement improves both gender equality and contraceptive success. The data also showed a strong statistical relationship (χ² = 49.423, p = 0.000), which supports the idea that shared responsibility is key in reproductive health.

Even though many men provide financial support for contraceptive methods, the study found that 60.2% of men do not go to clinics with their wives. This finding is similar to what(Mulatu et al., 2022) observed: while men often help financially, they are less involved emotionally or practically in family planning. Men in the study often talked about providing money but did not see the need to be involved in clinic visits or decision-making. This shows that more needs to be done to promote full involvement of men in family planning, not just financial support.

Most people in the study (98.6%) were aware of long-acting reversible contraceptives (LARC), showing high general awareness. This aligns with findings by (Yadassa et al., 2023), who said education and health campaigns help raise awareness about family planning. However, only 38.7% of respondents reported making LARC decisions together with their partners. This is similar to what(Tumwizere et al., 2024) found in sub-Saharan Africa while people know about contraceptives, joint decision-making is still low, especially in traditional communities.

The gap between knowing about LARC and making shared decisions could be due to gender roles and power differences. Even though both men and women may see LARC as useful, cultural norms may limit how much men are involved in decisions. Some women in the study said they chose their contraceptives alone, even though they knew they should talk to their partners. This matches what(Namasivayam et al., 2022) found that gender gaps in decision-making remain, despite greater awareness.

The findings in Figure 2 of this study align with existing literature that emphasizes the critical role of healthcare providers in disseminating accurate information about long-acting reversible contraceptives (LARCs). Similar to studies conducted in Uganda and other sub-Saharan African countries, healthcare workers were the most cited source of information (43.9%), reinforcing their influence on women’s contraceptive choices. The involvement of friends (18.5%) and family (21.5%) as key information sources reflects the importance of interpersonal communication in shaping health behavior, as noted by (Wambete et al., 2024), who observed that peer and family influence significantly affect contraceptive uptake. However, the low reliance on schools (11.3%) and media (4.7%) suggests a gap in formal and mass communication strategies, echoing findings from (Zandam et al., 2022), which recommended strengthening school-based and multimedia family planning education to improve awareness and promote male involvement in reproductive health.

Source: Field Survey 2025

**Figure 2:**
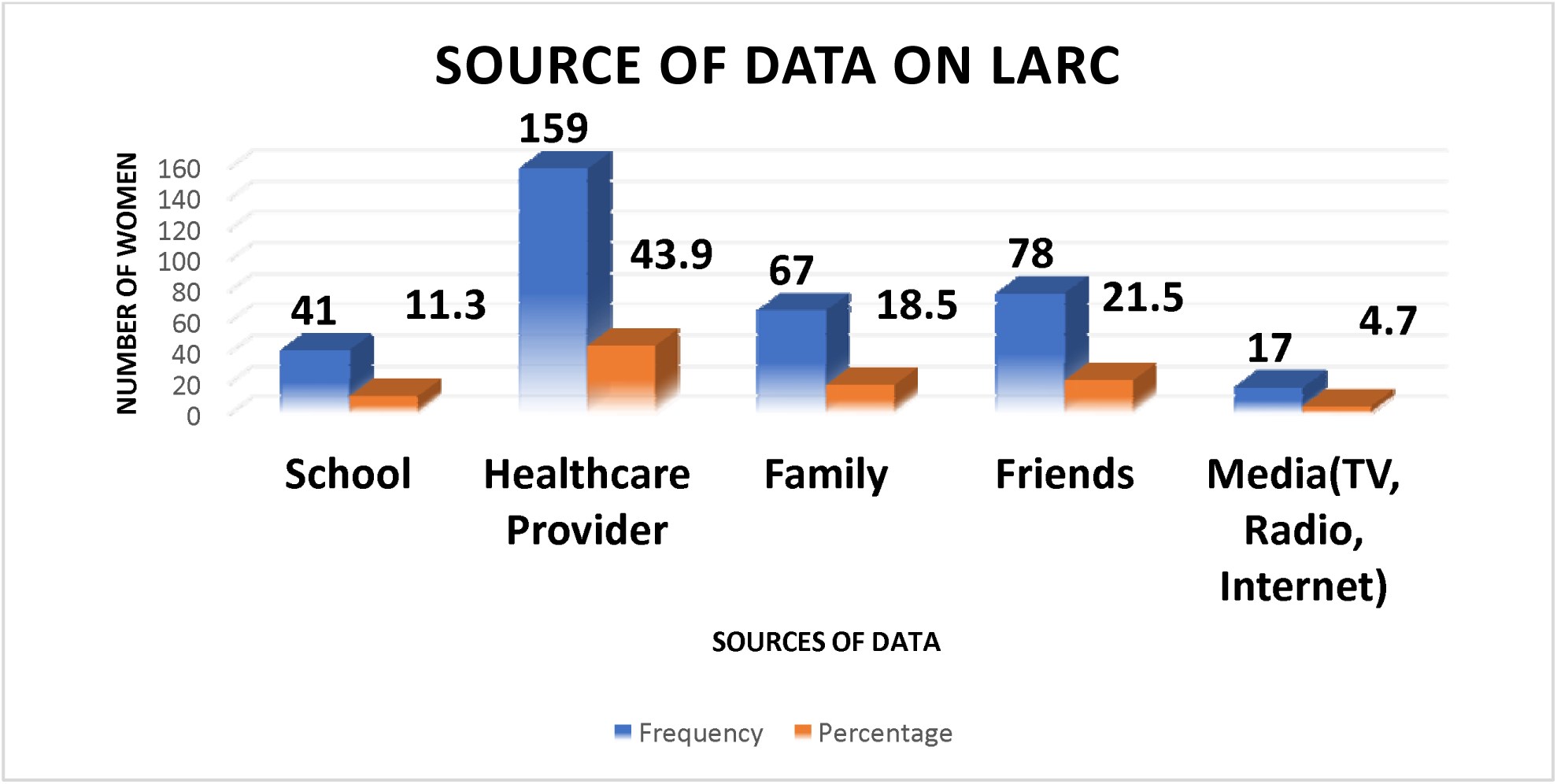
Source of Data on LARC. Male respondents reported learning about long-acting reversible contraceptive (LARC) methods from various sources. The majority (43.9%) said healthcare providers were their main source of information. Family members (21.5%) and friends (18.5%) also played key roles. Smaller numbers learned about LARCs through school (11.3%) or media platforms like TV, radio, and the internet (4.7%). These findings highlight the diverse channels through which LARC information is shared, which can influence women’s choices and the level of male involvement in family planning decisions, as shown in Figure (2) below. Source: Field Survey 2025

The findings from the Chi-square analysis Table 2 above provide important insights into the factors that influence male involvement in the use and decision-making surrounding long-acting reversible contraceptives (LARC) in Kampala, Uganda. Several key factors were identified as significantly associated with male involvement in family planning decisions, including marital status, education level, number of children, monthly income range, and polygamous status. These factors offer critical implications for improving male engagement in reproductive health, which is crucial for the successful adoption of LARC methods and other family planning strategies.

**Table 2:** Chi-Square on factors and barriers influencing male involvement in long-acting reversible contraceptives (LARC) use and decision-making in Kampala, Uganda.

| Variable | Response | Frequency (%) | $\chi^2$ | p-value |
| --- | --- | --- | --- | --- |
| **Marital Status** | Single | 22 (6.1%) | 35.627 | **0.001\*** |
|  | Married | 312 (86.2%) |  |  |
|  | Cohabiting | 16 (4.4%) |  |  |
|  | Divorced/Separated | 9 (2.5%) |  |  |
|  | Widowed | 3 (0.8%) |  |  |
|  | **Total** | **362 (100%)** |  |  |
| **Education Level** | No formal education | 8 (2.2%) | 44.289 | **0.002\*** |
|  | Primary school | 57 (15.7%) |  |  |
|  | Secondary school | 157 (43.4%) |  |  |
|  | Diploma/Certificate | 109 (30.1%) |  |  |
|  | Bachelor's degree | 24 (6.6%) |  |  |
|  | Master's degree or higher | 7 (1.9%) |  |  |
|  | **Total** | **362 (100%)** |  |  |
| **Number of Children** | None | 19 (5.2%) | 38.428 | **0.004\*** |
|  | One to Two | 140 (38.7%) |  |  |
|  | Three to Four | 174 (48.1%) |  |  |
|  | Five to Six | 28 (7.7%) |  |  |
|  | Seven or More | 1 (0.3%) |  |  |
|  | **Total** | **362 (100%)** |  |  |
| **What do you think prevents men from being more involved in LARC?** | Lack of knowledge | 12 (3.3%) | 21.053 | 0.636 |
|  | Cultural beliefs | 31 (8.6%) |  |  |
|  | Religious beliefs | 76 (21.1%) |  |  |
|  | Fear of side effects | 129 (35.7%) |  |  |
|  | Partner's preference | 108 (30.0%) |  |  |
|  | No barriers | 4 (1.1%) |  |  |
|  | **Total** | **361 (100%)** |  |  |
| **Cultural norms affect your involvement in LARC?** | Strongly disagree | 6 (1.7%) | 42.813 | **0.000\*** |
|  | Disagree | 15 (4.1%) |  |  |
|  | Neutral | 45 (12.4%) |  |  |
|  | Agree | 264 (73.0%) |  |  |
|  | Strongly agree | 32 (8.8%) |  |  |
|  | **Total** | **362 (100%)** |  |  |
| **Do religious beliefs influence your decisions about LARC** | Strongly disagree | 9 (2.5%) | 29.402 | **0.021\*** |
|  | Disagree | 50 (13.8%) |  |  |
|  | Neutral | 135 (37.2%) |  |  |
|  | Agree | 146 (40.3%) |  |  |
|  | Strongly agree | 22 (6.1%) |  |  |
|  | **Total** | **362 (100%)** |  |  |

Male participation in decision-making about long-acting reversible contraceptives (LARC) is strongly linked to marital status. Married men are more likely than single or cohabiting men to engage in reproductive health decisions, which aligns with findings from other regions such as sub-Saharan Africa, where formal marriages support shared decision-making on family planning(Demissie et al., 2022). Marriage provides a framework for communication and shared responsibility in contraception choices.

Education is also very important. Higher-educated men are more likely to participate in LARC decisions, particularly those who have finished secondary school. This confirms previous studies that found education has a favorable impact on men’s views and support for using contraceptives(Msovela et al., 2020). Therefore, raising awareness of reproductive health could encourage more men to use family planning.

Another factor influencing a man’s engagement is the number of children he has. Due to their desire to limit the size of their family and prevent unwanted pregnancies, men who have three or four children are more likely to be active in the contraceptive decision-making process. This result is consistent with earlier research conducted in Uganda, which found that as families expand, men’s involvement rises(Tumwizere et al., 2024).

However, men’s participation is restricted by a number of obstacles. Common barriers include partner choice (30.0%), cultural norms (8.6%), religious beliefs (21.1%), and side effect fears (35.7%). These elements align with research conducted in sub-Saharan Africa, where the use of contraceptives is frequently discouraged by cultural and religious considerations(Boadu, 2022). Many men are less supportive of their spouses’ usage of contraceptives because they are concerned about the possible negative health effects of LARC, such as weakness and excessive bleeding(Jonas et al., 2022).

Preferences of the partner are also important. Given the prevalence of women in Uganda making family planning decisions without consulting males, many men stated that their female partners frequently made decisions about contraception on their own(Muhumuza et al., 2023). This emphasizes the necessity of family planning initiatives that encourage collaborative decision-making in cultures with strong patriarchal norms.

Cultural and religious standards heavily influence male participation in LARC decisions. 40.3% of respondents acknowledged the importance of religion, while the majority (73%) believed that cultural norms have an impact on male engagement. Other research that highlights how cultural and religious influences influence family planning views in Uganda supports these findings(Anaman-Torgbor et al., 2025). Male involvement in reproductive health is sometimes limited by traditional gender roles, which place the burden on women. The use of contraceptives is also limited by religious objections, particularly in Muslim and Christian groups(Achen et al., 2021).

Finally, some participants said that talking about contraception helped their relationship. This agrees with (Ontiri et al., 2021), who also said communication is key. But these conversations are still rare, partly due to cultural taboos. This shows that there is a need to encourage more open conversations between partners. This could help improve family planning and relationship quality.

Qualitative results revealed social stigma against women who use family planning. When women use contraception, they are subjected to criticism and rumors, which may deter them from using it further. There are gaps in male-targeted education, as evidenced by the fact that although 52.5% of men said they knew enough about LARC, a sizable number disagreed or had no opinion(Kriel et al., 2023). Misconceptions and mistrust around contraceptives are exacerbated by the absence of targeted reproductive health education for men.

In Table 3, Healthcare providers play a key role in promoting male involvement in family planning, particularly regarding long-acting reversible contraceptives (LARCs). However, they encounter challenges in engaging men in these discussions. While 44.2% of participants believed that healthcare providers effectively address men’s concerns about LARCs, a significant portion disagreed or were undecided. The chi-square test indicated no significant difference (χ² = 17.663, p = 0.344), suggesting mixed experiences, likely stemming from gaps in communication and outreach(Kibonire & Mphuthi, 2023).

**Table 3:**
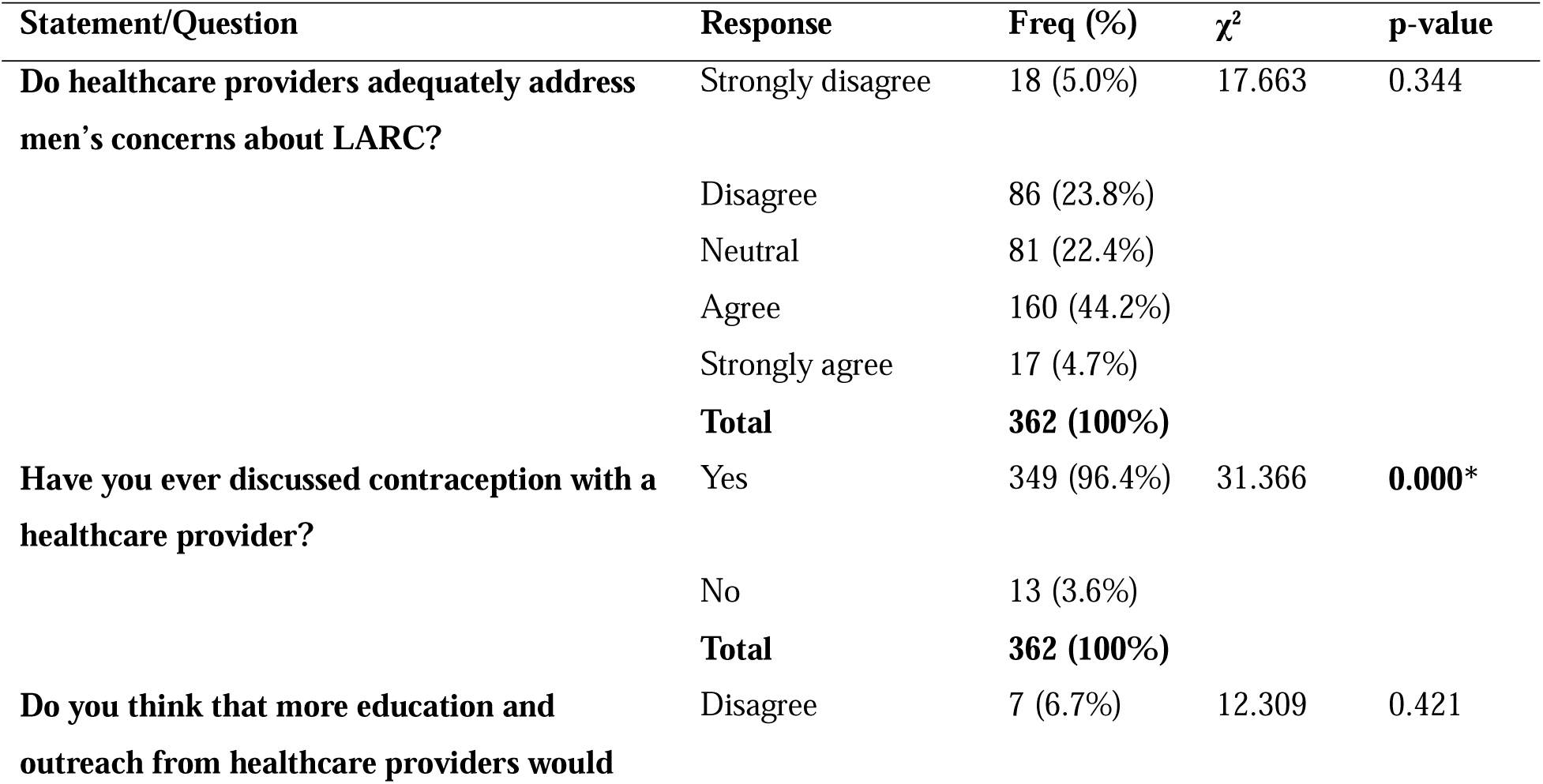
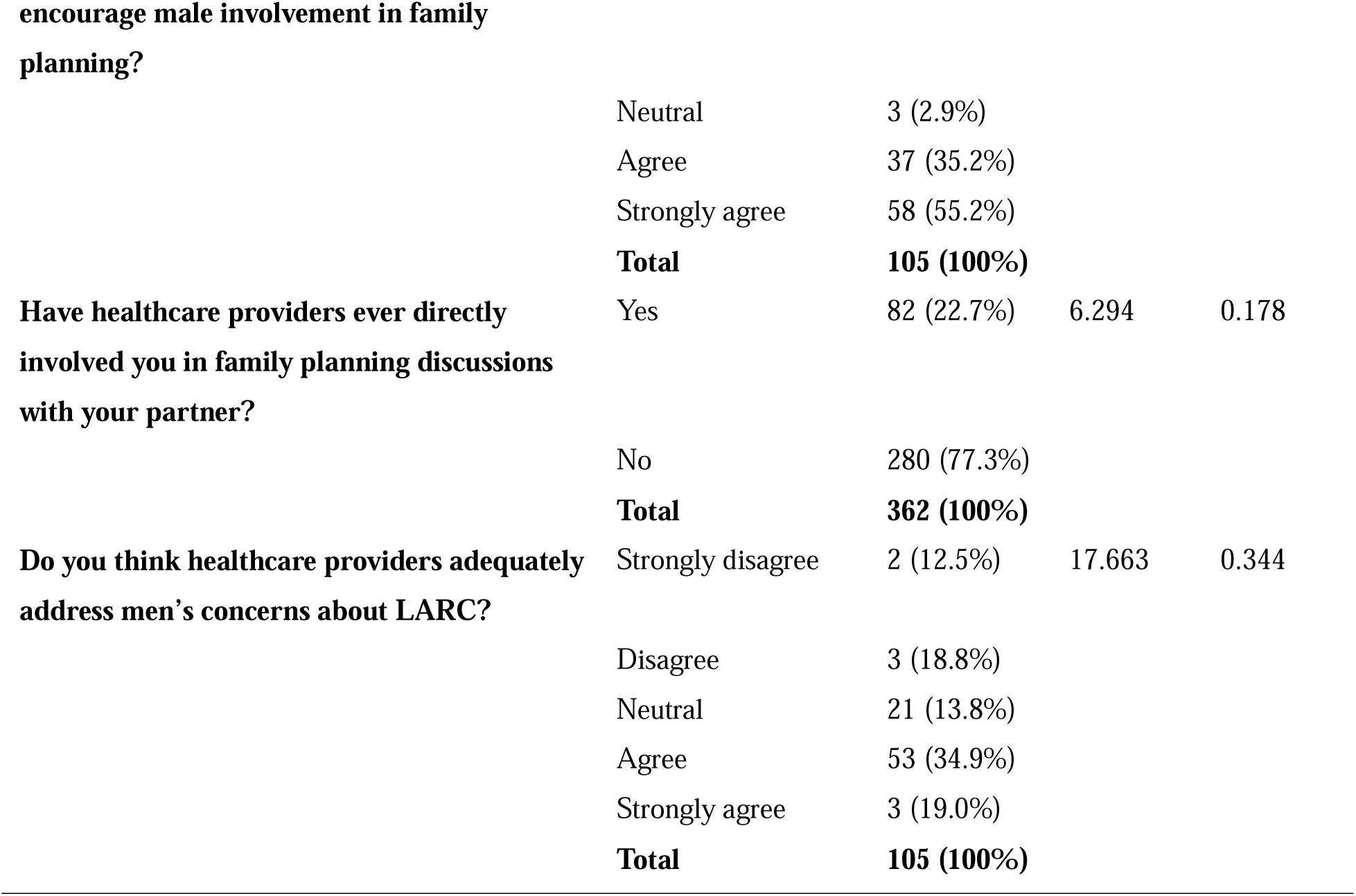
Chi-Square analysis on the role of healthcare providers in promoting male involvement in family planning, especially long-acting reversible contraceptives.

Most participants (96.4%) had spoken to a healthcare provider about contraception, and this result was statistically significant (χ² = 31.366, p = 0.000). However, family planning is still often treated as a woman’s issue, with men frequently left out of conversations. Involving both partners in counseling sessions could improve shared decision-making(Hernandez et al., 2022).

More provider education and outreach were strongly supported 35.2% agreed and 55.2% strongly agreed that it would assist improve male involvement. Results by (Wambete et al., 2024), who emphasized the significance of tailored education for men, are supported by this. Although there was no significant difference according to the chi-square test (χ² = 12.309, p = 0.421), this perception might be influenced by regional or cultural factors.

The fact that 77.3% of respondents claimed that their male partners had not been actively included in family planning conversations by healthcare practitioners was a worrying finding. This confirms the findings of (Schwandt et al., 2021), who discovered that health professionals frequently give preference to women while counseling, presuming that men are uninterested. The lack of a significant difference (χ² = 6.294, p = 0.178) indicates that this is a common problem that requires focused interventions to promote collaboration.

### Qualitative Insights and Integration with Quantitative Data

The survey results were corroborated by qualitative information gathered from focus groups and interviews, which also brought to light important concerns regarding male participation in family planning, particularly with LARCs. Many men claimed that because they did not have direct access to medical professionals, their husbands were hesitant to talk about family planning. “*I believe my husband would feel more at ease if the doctor spoke to him directly about LARC methods*,” one woman stated. Men were rarely involved, even though the majority of women spoke with providers about contraception. “*I’ve visited the clinic numerous times to talk about contraception, but my husband has never been involved,*” one lady revealed.

Both men and women stressed the need for joint education and counseling. A man said, *“If I had more information about contraception, I would take part more.”* A woman added, *“If my husband understood the options, he would be more supportive.”* Despite this, 77.3% said men were not included by providers in discussions.

There were several misconceptions regarding LARCs. According to a medical professional, “Many men are scared by the idea that these methods are not reversible.” *Male involvement is limited by this false idea. Nonetheless, there is growing acceptance for shared responsibility. “It’s important that men are involved*,” one woman stated. “*It’s my responsibility to understand contraception*,” a male concurred.

Male active engagement is minimal, despite great awareness of LARCs. As one man acknowledged, “*I provide money… but I don’t go to the clinic*,” many men offer financial support but do not accompany partners to clinics. This is consistent with survey results that indicate men’s involvement is limited to financial matters.

### Limitations

This study may not accurately represent the opinions of the general public because it was restricted to men who were enrolled at a particular wellness clinic in Kampala’s Makindye division. There was no specific survey for women; instead, FGDs were used to gather women viewpoints indirectly. Furthermore, recall or social desirability bias might have been introduced by self-reported replies. Finally, causal conclusions between variables are not possible due to the cross-sectional design.

## Supporting information

Supplemental Questionaire

## Data Availability

The datasets generated and/or analyzed during the current study are available from the corresponding author upon reasonable request. Due to the sensitive nature of the information collected, all data have been anonymized to protect participants' identities.

## Ethics Approval and Consent to Participate

The Uganda National Council for Science and Technology (UNCST) granted ethical approval for this work under approval number HS5778ES, and the Kampala International University Research Ethics Committee (KIU-REC) granted ethical approval under approval number KIU-2024-563. The study was conducted in accordance with the ethical principles outlined in the Declaration of Helsinki. Before participating in the trial, each subject gave written informed consent. Anonymity and confidentiality were maintained during the data collection and reporting procedures, and participation was entirely voluntary.

## Consent for Publication

Not applicable. This manuscript does not contain any individual participant data such as images, videos, or identifiable personal information requiring consent for publication.

## Authors’ Contributions

Brian Nyasulu conceptualized the study, conducted fieldwork, performed data analysis, and drafted the manuscript. Dr. Nicholas Ngomi, Dr Anumolu Goparaju and Dr. Ronald Arineitwe Kibonire provided academic supervision, reviewed the methodology, and contributed to the critical revision of the manuscript. All authors read and approved the final manuscript.

## Authors’ Information (Optional)

Brian Nyasulu is a postgraduate student in the Department of Public Health at Kampala International University.

Dr. Nicholas Ngomi, Dr Anumolu Goparaju and Dr. Ronald Arineitwe Kibonire are faculty members in the Department of Public Health, Kampala International University, specializing in reproductive health and health policy, respectively.

## Conclusion

This study provides important new information about the variables influencing men’s participation in long-acting reversible contraceptive (LARC) decision-making in Kampala, Uganda. The results show that male involvement is primarily restricted to offering financial assistance, despite the fact that there is a high level of awareness about LARC methods and many people think using contraceptives improves relationships. This disparity highlights the necessity of defining male involvement more broadly to encompass active decision-making, practical help, and emotional support in addition to monetary contributions. Income, the number of children, education, marital status, and polygamous partnerships all have a significant impact on men’s participation in LARC choices. Generally speaking, family planning is more common among males in stable marriages who are better educated, have more children, and earn more money. However, active engagement is frequently limited by misconceptions regarding partner preferences, side effects, and traditional gender roles. Religious and cultural convictions can provide serious obstacles to male participation. These difficulties, along with the lack of candid communication between partners over family planning, underscore the need of initiatives that encourage open communication and tackle societal and cultural norms that restrict men’s use of contraceptives.

