## Supplemental Questionaire for "EXPLORING MALE INVOLVEMENT IN CONTRACEPTIVE DECISION-MAKING AND SUPPORT FOR LONG-ACTING REVERSIBLE CONTRACEPTIVES AMONG COUPLES IN KAMPALA, UGANDA"

### QUESTIONNAIRE

#### Introduction

My name is Brian Nyasulu, and I am currently pursuing a Master's degree in Public Health at Kampala International University. As part of my academic research, I am conducting a study on **Male Involvement in Contraceptive decision-making and Support for Long-Acting Reversible Methods among Couples in Kampala, Uganda**. The purpose of this study is to explore the roles, perceptions, and attitudes of men regarding contraceptive use and how these factors influence decision-making in relationships.

Your participation in this survey is entirely voluntary, and all information you provide will be kept strictly confidential. The data collected will be used solely for academic purposes and will help in understanding and promoting male involvement in family planning initiatives.

Thank you for taking the time to participate in this important study. Your insights are invaluable and will contribute significantly to the success of this research.

#### Section 1: Demographic Information

1. **What is your age?**
    - Under 20
    - 21-30
    - 31-40
    - 41-50
    - 51 and above
2. **What is your highest level of education?**
    - No formal education
    - Primary school
    - Secondary school
    - Diploma/Certificate
    - Bachelor's degree

- Master’s degree or higher

3. **What is your marital status?**
   - Single
   - Married
   - Cohabiting
   - Divorced/Separated
   - Widowed
4. **How long have you been in your current relationship?**
   - Less than 1 year
   - 1-3 years
   - 4-7 years
   - 8-10 years
   - More than 10 years
5. **How many children do you have?**
   - None
   - 1-2
   - 3-4
   - 5-6
   - 7 or more
6. **What is your occupation?**
   - Unemployed
   - Self-employed
   - Employed in the private sector
   - Employed in the public sector
   - Student
   - Other (please specify): ____________
7. **What is your monthly income range?**
   - Less than UGX 100,000
   - UGX 100,000 - 300,000
   - UGX 300,001 - 500,000
   - UGX 500,001 - 1,000,000

- More than UGX 1,000,000

8. **Which religious group do you belong to?**
   - Christian (non-denominational)
   - Roman Catholic
   - Muslim
   - Anglican
   - Pentecostal
   - Other (please specify): ____________
9. **Are you the only wife to your husband?**
   - Yes
   - No

**Section 2: Knowledge and Attitudes towards Contraception**

10. **Have you heard about contraceptive methods?**
    - Yes
    - No
11. **Which contraceptive methods are you familiar with?** (Select all that apply)
    - Birth control pills
    - Injections
    - Implants
    - Intrauterine device (IUD)
    - Natural methods (e.g., withdrawal)
    - Other (please specify): ____________
12. **Where did you first learn about contraception?**
    - School
    - Healthcare provider
    - Family
    - Friends
    - Media (TV, radio, internet)
    - Other (please specify): ____________
13. **Do you believe that using contraceptives is important for family planning?**

- Strongly agree
- Agree
- Neutral
- Disagree
- Strongly disagree

14. **What is your perception of the safety of contraceptive implants?**
    - Very safe
    - Safe
    - Neutral
    - Unsafe
    - Very unsafe
15. **Do you think that men should be involved in contraceptive decision-making?**
    - Strongly agree
    - Agree
    - Neutral
    - Disagree
    - Strongly disagree
16. **Do you believe that contraceptive use can improve a couple's relationship?**
    - Strongly agree
    - Agree
    - Neutral
    - Disagree
    - Strongly disagree
17. **What are your thoughts on the long-acting effects of contraceptive use on women's health?**
    - Very positive
    - Positive
    - Neutral
    - Negative
    - Very negative

**Section 3: Involvement in Contraceptive Decision-Making**

18. **Who usually makes decisions about contraception in your relationship?**
    - Myself
    - My partner
    - Both of us equally
19. **How often do you discuss contraceptive options with your partner?**
    - Always
    - Often
    - Sometimes
    - Rarely
    - Never
20. **Have you ever participated in choosing a contraceptive method with your partner?**
    - Yes
    - No
21. **Do you feel comfortable discussing contraception with your partner?**
    - Very comfortable
    - Comfortable
    - Neutral
    - Uncomfortable
    - Very uncomfortable
22. **Have you ever been discouraged by your partner from using any contraceptive method?**
    - Yes
    - No
23. **If yes, why does he discourage you from the use of that contraceptive method?** (Open-ended question)
24. **Would you support your partner's decision to use long-acting contraceptive methods (e.g., implants)?**
    - Strongly agree

- Agree
- Neutral
- Disagree
- Strongly disagree

25. **Do you think using contraception can have positive effects on your relationship?**
    - Strongly agree
    - Agree
    - Neutral
    - Disagree
    - Strongly disagree
26. **Do you think your community supports male involvement in family planning?**
    - Strongly agree
    - Agree
    - Neutral
    - Disagree
    - Strongly disagree
27. **How involved do you believe men should be in family planning?**
    - Very involved
    - Involved
    - Neutral
    - Minimally involved
    - Not involved at all
28. **How does your husband support you in the choice of contraceptive method?** (Select all that apply)
    - Accompany her to the clinic and discuss the options with her
    - Provide financial support but do not accompany me to the Hospital
    - Offer emotional support but do not accompany me to the Hospital
    - Do not support
    - Other (please specify): ____________

**Section 4: Barriers to Male Involvement**

29. **What prevents your husband from being more involved in contraceptive decision-making?** (Select all that apply)
    - Lack of knowledge
    - Cultural beliefs
    - Religious beliefs
    - Fear of side effects
    - Partner’s preference
    - No barriers
    - Other (please specify): ____________
30. **Do you believe that contraceptives can negatively affect a woman’s health?**
    - Strongly agree
    - Agree
    - Neutral
    - Disagree
    - Strongly disagree
31. **How do you feel about the perception that family planning is primarily a woman’s responsibility?**
    - Strongly agree
    - Agree
    - Neutral
    - Disagree
    - Strongly disagree
32. **Do you think there is adequate information available to men about contraception?**
    - Strongly agree
    - Agree
    - Neutral
    - Disagree
    - Strongly disagree

33. **Do you face any stigma in your community for being involved in family planning?**
    - o Yes
    - o No
34. **If yes, what type of stigma do you experience?** (Open-ended question)
35. **Would you like to receive more information about family planning methods?**
    - o Yes
    - o No
36. **Do you feel that cultural norms in your community affect your involvement in contraceptive decisions?**
    - o Strongly agree
    - o Agree
    - o Neutral
    - o Disagree
    - o Strongly disagree
37. **Do religious beliefs influence your decisions about contraceptive use?**
    - o Strongly agree
    - o Agree
    - o Neutral
    - o Disagree
    - o Strongly disagree
38. **Have you experienced any societal pressure regarding contraceptive use?**
    - o Yes
    - o No
39. **If yes, what kind of societal pressure have you experienced?** (Select all that apply)
    - o Pressure to avoid contraceptives
    - o Pressure to use contraceptives
    - o Pressure to have more children
    - o Other (please specify): ____________

**Section 5: Role of Healthcare Providers**

40. **Have you ever discussed contraception with a healthcare provider?**

- Yes
- No

41. **If yes, how satisfied were you with the information provided?**
    - Very satisfied
    - Satisfied
    - Neutral
    - Dissatisfied
    - Very dissatisfied
42. **Do you think healthcare providers adequately address men's concerns about contraception?**
    - Strongly agree
    - Agree
    - Neutral
    - Disagree
    - Strongly disagree
43. **Have healthcare providers ever directly involved you in family planning discussions with your partner?**
    - Yes
    - No
44. **Do you believe that more education and outreach from healthcare providers would encourage male involvement in family planning?**
    - Strongly agree
    - Agree
    - Neutral
    - Disagree
    - Strongly disagree

Conclusion

Thank you for completing this questionnaire. Your responses will significantly contribute to understanding the dynamics of contraceptive decision-making in couples and promote male involvement in family planning initiatives. If you have any additional comments or suggestions regarding this study, please feel free to share them below:

#### Semi-structured interview Questions on FGDs

My name is Brian Nyasulu, and I am currently pursuing a Master’s degree in Public Health at Kampala International University. As part of my academic research, I am conducting a study titled:

"Male Involvement in Contraceptive Decision-Making and Support for Long-Acting Reversible Methods among Couples in Kampala, Uganda."

This discussion is aimed at exploring the roles, perceptions, experiences, and attitudes of both men and women regarding family planning—particularly around long-acting reversible contraceptive methods, such as implants and IUDs—and how men’s involvement influences decision-making within couples.

We believe that understanding both male and female perspectives is key to strengthening family planning programs and promoting shared responsibility in reproductive health.

Your participation is completely voluntary, and you are free to express your views openly. Please know that everything shared here will be treated with strict confidentiality and will be used solely for academic and research purposes. There are no right or wrong answers—we are interested in your honest opinions and lived experiences.

We encourage open and respectful conversation. Please allow each other to speak, and feel free to add to what others share. This is a safe space, and your input is highly valued.

Once again, thank you for taking the time to participate. Your voices are crucial in helping us understand how to better involve men in family planning and support healthier, more informed decisions among couples**.**

**Introduction**

1. Understanding Perceptions and Attitudes

- What are your thoughts on the use of contraceptives in your community?
- How do you feel about men being involved in decisions about contraception?

2. Cultural and Religious Influences

- In what ways do cultural practices influence men's participation in family planning?
- How do religious beliefs shape your views on contraceptive use?

3. Decision-Making Processes

- Who usually takes the lead in making decisions about contraception in your household?
- How do you and your partner discuss and decide on contraceptive methods?

4. Barriers to Male Involvement

- What challenges do you face in participating in contraceptive decisions?
- Can you identify any societal or personal barriers that prevent men from getting involved in family planning?

5. Communication with Partners

- How open are you with your partner when discussing family planning and contraception?
- What are the most effective ways you've found to communicate about contraceptives with your partner?

6. Experiences with Healthcare Providers

- What has your experience been like when talking to healthcare providers about contraception?
- Do you feel healthcare providers encourage male involvement in contraceptive decisions?

7. Community and Social Support

- How does the community view men who are involved in family planning?
- What kinds of support or resources do you think would encourage more men to participate in contraceptive decisions?

8. Recommendations and Future Directions

• What changes would you recommend to increase male involvement in contraceptive decision-making?

• How can the community better support men in taking an active role in family planning?

**Questions for KII Healthcare Workers**

1. Training and Awareness

o What kind of training have you received on engaging men in family planning, especially in promoting long-acting reversible contraceptive methods?

o How prepared do you feel to address men's concerns and questions about long-acting reversible contraceptives?

2. Patient Engagement

o How do you involve men in family planning discussions when couples come in for consultations?

o What strategies do you use to encourage male participation in choosing long-acting reversible contraceptive methods?

3. Barriers and Challenges

o What challenges do you face when trying to promote male involvement in family planning, particularly with long-acting reversible contraceptives?

o Can you share any common misconceptions or fears that men have about long-acting reversible contraceptive methods?

4. Improvement and Support

o What additional support or resources would help you better promote male involvement in family planning?

o How do you think healthcare services could be improved to make men more comfortable and active in family planning decisions, especially concerning long-acting reversible contraceptives?
